# Beat-to-beat blood pressure variability and Alzheimer’s disease pathophysiology

**DOI:** 10.64898/2026.11.10.26367476

**Authors:** Trevor Lohman, Jillian Joyce, Melanie Quiring, Basant I. Lashin, Arunima Kapoor, Allison C Engstrom, Kevin W. Law, Farrah Ferrer, Lorena Contreras, John Paul M. Alitin, Aimee Gaubert, Kathleen E Rodgers, Elizabeth Head, Rond Malhas, Niema Pahlevan, Daniel A Nation

## Abstract

**Introduction:** Elevated blood pressure variability (BPV) is predictive of Alzheimer’s disease neuropathologic change observed with AD biomarkers. However, prior studies have largely focused on visit-to-visit BPV, which may be influenced by antihypertensive medication adherence and other external factors. Few studies have investigated beat-to-beat BPV, a reliable and direct measure of cardiovascular and autonomic function relevant to AD.

**Methods:** Independently living older adults (N=132) were recruited from the community. Participants underwent continuous blood pressure monitoring and venipuncture. Plasma p- Tau_217_, p-Tau_181_, GFAP, NfL, and amyloid-β_42/40_ ratio were assessed. Linear regression examined the relationship between beat-to-beat systolic BPV and plasma AD biomarkers with and without adjustment for age, sex, systolic blood pressure, and apolipoprotein E4 (*APOE4*) carrier status.

**Results:** Elevated beat-to-beat BPV was associated with elevated plasma p-Tau_217_ (β=0.32, p=0.008), plasma p-Tau_181_ (β=0.22, p=0.05), and plasma GFAP (β=0.41, p=0.00001). Elevated BPV was not associated with plasma amyloid-β_42/40_ ratio or plasma NfL.

**Discussion:** Increased beat-to-beat BPV is associated with increased Core 1 T_1_ biomarkers of phosphorylated and secreted AD tau in blood plasma (i.e., pTau_217_, pTau_181_), but not with amyloid-β_42/40_ ratio, a biomarker more exclusively indexing cerebral amyloid retention alone.

Elevated BPV was also associated with higher plasma GFAP, a marker of astrocyte reactivity. These findings are consistent with prior studies relating visit-to-visit BPV to AD biomarkers, suggesting that intrinsic cardiovascular and/or autonomic factors underlie links between BPV and AD.

## Introduction

Blood pressure variability (BPV) over various time scales is associated with cerebrovascular disease^1-7^, neurodegenerative disease^8,9^, cognitive decline^10-12^, and dementia^13-19^. Visit-to-visit BPV is a widely used convenience measure of BPV that is associated with accelerated Alzheimer’s disease (AD) neuropathologic change reflected in AD biofluid markers and molecular imaging^15,19-23^. However, questions remain regarding the pathogenesis of visit-to- visit BPV, its test-retest reliability, and the specific mechanisms linking high visit-to-visit BPV to AD pathophysiology. Blood pressure may vary over months and years between clinic visits due to numerous extraneous factors, including antihypertensive medication adherence and timing, seasonal and diurnal oscillations, temperature, diet and meal timing, psychological stress, and other factors^24^. Thus, it remains unclear whether the relationship between visit-to-visit BPV and AD biomarkers is the product of intrinsic cardiovascular and autonomic factors relevant to AD pathophysiology versus the many extraneous sources of BPV across visits.

Alternatively, beat-to-beat BPV is a valid^25,26^, high temporal resolution marker of cardiovascular and autonomic function and arterial stiffness^27^ that can be acquired reliably^28^ in a single clinic visit. Only a few small studies have investigated the relationship between beat-to- beat BPV and AD biomarker levels^10,29^, and these studies have not included plasma p-Tau_217_, a more sensitive and specific marker of amyloid and AD-tau pathophysiology^30-32^. Prior studies suggest that BPV, particularly fluctuations in systolic blood pressure between consecutive cardiac cycles, is more closely associated with AD-tau than with amyloid-β^10,20^. Therefore, the present study will test the a priori hypotheses that higher systolic beat-to-beat BPV is associated with increased plasma p-Tau_217_ and increased plasma glial fibrillary acidic protein (GFAP), two of the earliest biomarkers of preclinical AD pathophysiology^33-39^, in order to address current knowledge gaps regarding intrinsic cardiovascular function and its effects on AD pathophysiology.

## Methods

### Participants

Participants were recruited from Los Angeles County and Orange County communities through outreach events, mailing lists, word-of-mouth, online portals, a research volunteer registry, and through the local Alzheimer’s Disease Research Center (ADRC). All procedures were conducted as part of the Vascular Senescence and Cognition (VaSC) Study at the University of Southern California (USC) and the University of California Irvine (UCI). Older adults aged 60 to 89 years who were living independently were included (Table 1). Study exclusions were a prior diagnosis of dementia, history of clinical stroke, family history of dominantly inherited neurodegenerative disorders, current neurological or major psychiatric disorders that may impact cognitive function, history of moderate-to-severe traumatic brain injury, active substance abuse, current use of medications impairing the central nervous system, current organ failure or other uncontrolled systemic illness, and contraindications for brain MRI. Eligibility for the study was verified by a structured clinical health interview and review of current medications with the participant and, when available, a knowledgeable informant study partner. This study was approved by the USC and UCI Institutional Review Board, and all participants gave informed consent. The anonymized data that support the findings of this study are available upon reasonable request from the corresponding author, DAN, through appropriate data sharing protocols.

| **Variable Name** | **Mean±SD** |
|---|---|
| Age (years) | 69.73±7.25 |
| Sex (n female, (%)) | 85 (64) |
| Systolic blood pressure (mmHg) | 132.55±18.77 |
| Systolic blood pressure average real variability (mmHg) | 1.92±1.03 |
| Taking antihypertensive medication (n, (%)) | 23 (17.42) |
| APOE Genotype |  |
| 2/2 (n, (%)) | 0 (0.0) |
| 2/3 (n, (%)) | 3 (2.3) |
| 3/3 (n, (%)) | 68 (51.5) |
| 2/4 (n, (%)) | 1 (0.8) |
| 3/4 (n, (%)) | 53 (40.2) |
| 4/4 (n, (%)) | 3 (2.3) |
| Missing (n, (%)) | 4 (3.0) |
| Plasma p-Tau<sub>217</sub> (pg/ml) | 0.35±0.23 |
| Plasma p-Tau<sub>181</sub> (pg/ml) | 25.14±10.53 |
| Plasma neurofilament light chain (pg/ml) | 16.21±6.04 |
| Plasma glial fibrillary acidic protein (pg/ml) | 145.91±79.06 |
| Plasma Aβ<sub>42/40</sub> ratio | 0.037±0.008 |

### Continuous BP data acquisition

Participants were asked to take medications as normally prescribed and abstain from caffeine on the morning of data collection. Beat-to-beat blood pressure measurements were obtained continuously during supine rest in a 3 T Siemens MRI scanner, using an MRI- compatible non-invasive continuous BP finger cuff device (Biopac®). First, the participant rests for 3 min in the supine position prior to the calibration period. During calibration, BP waveforms are acquired by the continuous monitoring device and two static pressures are simultaneously acquired using a calibrated, MRI-compatible automatic BP device with an inflatable brachial artery cuff (TeslaDUO). These static pressures are used to calibrate the continuous BP monitor using the Caretaker® system (Biopac®). After calibration, continuous BP was monitored during MRI for 7 min and further data processing was performed as previously described^28^.

### Systolic blood pressure average real variability

Beat-to-beat BPV is quantified as systolic blood pressure (SBP) average real variability (ARV), a measure of systolic beat-to-beat BPV which has been demonstrated as reliable in older adults^28,40^ regardless of antihypertensive medication use^28^. ARV calculates the average of absolute changes between consecutive blood pressure readings and is calculated as:

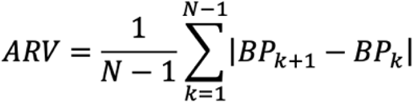

where n represents the number of blood pressure readings obtained during continuous blood pressure monitoring and k represents the beat index of the readings as previously described^28,40^. Systolic ARV was chosen as the measure of beat-to-beat BPV in the present study due to its increased reliability compared to other BPV measures such as standard deviation and coefficient of variation^28^, and its decreased susceptibility to outliers^40^. Additionally, ARV has the advantage of considering the temporal ordering of systolic BP measurements and is therefore a more specific measure of beat-to-beat fluctuations in blood pressure^28,41^.

### APOE genotyping

Apolipoprotein E genotype was determined from blood-derived genomic DNA using one of two methods: TaqMan SNP genotyping assays (rs429358 and rs7412; Thermo Fisher Scientific) or a PCR–restriction fragment length polymorphism protocol with HhaI digestion, as previously described^42^. *APOE4* carriers were defined as having at least one ε4 allele and coded as a binary variable (carrier = 1, non-carrier = 0) for analyses.

### Plasma biomarkers

Plasma p-Tau_217_ levels were obtained using the ALZpath p-Tau 217 Advantage PLUS kit (Quanterix), following the manufacturer’s protocol.Plasma levels of Aβ_40_ and Aβ_42_ were determined, and the Aβ_42/40_ ratio was calculated as an index of cerebral amyloid retention^43^. Plasma levels of Aβ_40_ and Aβ_42_ were obtained by digital immunoassay, Simoa Neurology 3-Plex A Advantage Kit (Quanterix), following the manufacturer’s protocol. Accepted ranges were as follows: Aβ_40_= 0–560 pg/mL, Aβ_42_= 0–240 pg/mL. Plasma pTau _**181**_ levels were obtained by the Simoa p-Tau_181_ Advantage V2.0 and 2.1 Assay (Quanterix), following the manufacturer’s protocol. Accepted ranges were 8–1,280 pg/mL. Plasma levels of GFAP and NfL were determined using Simoa Neurology 2-Plex B (N2PB) Kit (Quanterix), following the manufacturer’s protocol on the HD-X machine. Accepted ranges were as follows: NfL□= □0– □∼□2000 pg/mL and GFAP□= □0– □∼□40,000 pg/mL. All biomarker assays were conducted in the same lab at UCI (EH).

### Data analysis

One hundred and thirty-six participants underwent continuous BP monitoring and plasma AD biomarker assay. Two outliers with abnormally high BPV (+3.44 and +7.96 SD), and two outliers with abnormally high p-Tau_217_ (+3.21 and +4.34 SD) were removed resulting in a total analyzed sample size of 132 participants. One participant was also missing plasma GFAP.

Linear regression models assessed whether beat-to-beat BPV was associated with plasma AD biomarkers with and without adjustment for age, sex, systolic blood pressure (SBP), *APOE4* carrier status, and plasma Aβ_42/40_ ratio (where applicable). All analyses were performed in R^44^.

## Results

132 participants were included in the present analysis, their characteristics and demographic data are reported in **Table 1**.

In univariate linear regression analysis, elevated beat-to-beat BPV was associated with elevated plasma p-Tau_217_ (β=0.32, p=0.008), plasma p-Tau_181_ (β=0.22, p=0.05), and plasma GFAP (β=0.41, p=0.00001). After adjustment for age, sex, systolic blood pressure, *APOE4* carrier status, and plasma amyloid-β_42/40_ ratio, elevated BPV remained significantly associated with p-Tau_217_ (β=0.36, p=0.008), plasma p-Tau_181_ (β=0.26, p=0.047), and plasma GFAP (β=0.38, p=0.0001). Elevated BPV was not associated with plasma amyloid-β_42/40_ ratio in the univariate model or in the model adjusted for age, sex, systolic blood pressure, and *APOE4* carrier status. BPV was also not associated with plasma NfL in either the univariate or adjusted model. Selected results are shown in **Figure 1**.

**Figure 1:**
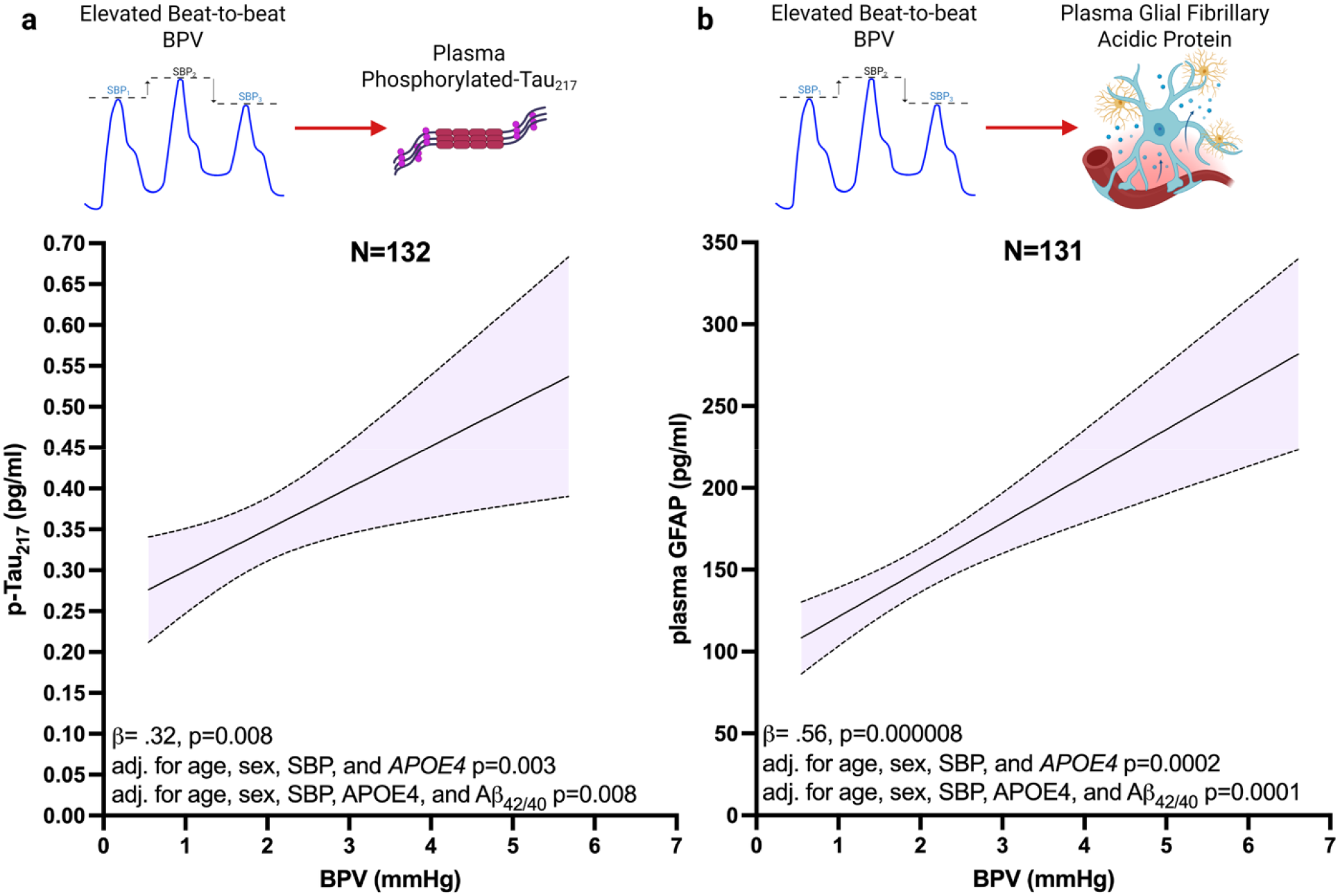
The relationship between beat-to-beat systolic blood pressure average real variability (BPV) and plasma p-Tau_217_, glial fibrillary acidic protein (GFAP), amyloid-β_42/40_ ratio, and neurofilament light chain. Standardized β-coefficients and p-values reported for significant univariate relationships followed by p-values for additional models with covariate adjustment. The relationship between elevated beat-to-beat BPV and plasma p-Tau_217_ is displayed in panel a, and plasma GFAP is shown in panel b. Plotted regression lines are unadjusted and shaded regions indicate 95% confidence intervals.

### Sensitivity analyses

All analyses were repeated without excluding >3SD outliers. The effect of BPV on plasma p-Tau_217_ remained significant (β=0.41, p=0.0007), as did its effect on plasma p-Tau_181_ (β=0.41, p=0.002) and plasma GFAP (β=0.24, p=0.003), and covariate adjustment did not attenuate the findings. Additionally, the analyses were repeated excluding participants with an *APOE2* allele, which did not affect the reported findings, nor did adjustment for antihypertensive medication use. The p-Tau_181_ finding did not survive Benjamini–Hochberg false discovery rate (FDR) correction^45^ and should therefore be interpreted with caution.

## Discussion

The present study finds that elevated beat-to-beat BPV is associated with increased plasma p-Tau_217_ and plasma GFAP, independent of age, sex, systolic blood pressure, *APOE4* carrier status, and plasma amyloid-β_42/40_ ratio. Importantly, this is the first study to investigate the relationship between valid^25,26,46^ and reliable^28^ beat-to-beat BPV and plasma p-Tau_217_, a highly sensitive and specific diagnostic and prognostic marker of AD pathophysiology^30-32^.

Elevated beat-to-beat BPV was not associated with amyloid-β_42/40_ and remained associated with p-Tau_217_ even after adjustment for amyloid-β_42/40_ ratio, confirming previous suggestions^20^ that BPV is differentially related to AD-tau (Core 1 T_1_ biomarkers^31^) compared to amyloid pathophysiology (Core 1 A_1_ biomarkers^31^). This is consistent with prior findings that arterial stiffness and other vascular risk factors are associated with AD-tau but not amyloid^47-50^. Pulse pressure for example was previously found to be associated with phosphorylated tau across a broad age range, while it was only associated with cerebral amyloidosis in the oldest individuals and those with more advanced neurodegeneration^47,48^. Similar relationships between vascular risk factors and their differential effects on AD-tau versus amyloid have been observed with arterial stiffness^49^, hypertension^50^, and Framingham risk score^50^. The consistent pattern between various vascular risk factors and AD-tau pathophysiology may suggest a common mechanism. Both blood pressure variability and pulse pressure are associated with decreased cerebral perfusion for example, particularly in vulnerable medial temporal regions^2-4,51^.

The strong relationship between BPV and plasma GFAP observed in the present study may offer additional insight. Astrocytes are exquisitely sensitive to cerebrovascular pressure changes, acting as intracranial baroreceptors^52-55^. Sudden changes in cerebral perfusion pressure may trigger astrocyte reactivity^52,53,56-60^, and prior studies show that astrocyte reactivity is a key mechanism component of tau hyperphosphorylation in AD^35,39,61,62^. BPV may predispose astrocytes to a reactive phenotype, as its relationship with plasma GFAP suggests, which merits further study.

Limitations include the cross-sectional nature of the study, which limits causal inference. Strengths include the adjustment for numerous relevant covariates, including the primary genetic risk factor for AD, *APOE4*^*63*^, and plasma amyloid-β_42/40_ ratio where applicable. Future longitudinal studies are needed to confirm the temporal relationships between BPV and AD-tau. Importantly, the authors recognize that both amyloid-β_42/40_ and plasma p-Tau_217_ closely track amyloid deposition, however plasma amyloid-β_42/40_ generally plateaus relatively early in the AD continuum, while the sharpest upward trajectory and dynamic range of plasma p-Tau_217_ tightly parallel the widespread distribution of concurrent amyloid and tau pathophysiology^64-68^. Plasma p-Tau_217_ is a measure of tau hyperphosphorylation that occurs in the presence of amyloid-β^65,69^ a unique feature of AD progression, as reflected in the Core 1 T1 classification of p-Tau_217_ in the most recent recommendations for the diagnosis and staging of AD^31^. Therefore, while both plasma amyloid-β_42/40_ and plasma p-Tau_217_ track amyloid-β deposition, the biomarkers represent clinically distinct states of AD progression and are therefore well-suited for testing the effects of elevated BPV on AD pathophysiology.

The present study findings are highly consistent with prior research, including the observed relationships with Core 1T_1_^22^ , and the relationship with plasma GFAP^10^. Importantly however, the present study adds the novel and clinically significant finding that elevated beat-to- beat BPV is associated with plasma p-Tau_217_ independent of blood pressure level which suggests BPV is a distinct vascular risk factor for AD, and that beat-to-beat BPV screening may be an important non-invasive early biomarker of Alzheimer’s disease.

## Data Availability

Data used in this article were obtained from the Global Parkinson Genetics Program (GP2; https://gp2.org). We used TIER 2 data from GP2 Release 11 (https://doi.org/10.5281/zenodo.17753486). GP2 data access can be requested through AMP PD (https://amp-pd.org). PPMI data used in this article were obtained on 26th August 2025 from the PPMI database (www.ppmi-info.org/access-data-specimens/download-data), RRID: SCR_006431. For information on the study, visit www.ppmi-info.org.
Sequencing data generated with ON and LN cannot be shared publicly, as participant consent does not cover public data sharing. Participant-level GBA1 calls from reference sequencing and NBA for all participants are provided in Supplementary Data 1.

## References

1. Sible IJ, Bangen KJ, Blanken AE, Ho JK, Nation DA. Antemortem Visit-To-Visit Blood Pressure Variability Predicts Cerebrovascular Lesion Burden in Autopsy-Confirmed Alzheimer’s Disease. Journal of Alzheimer’s Disease. 2021;83:65–75. doi:10.3233/JAD-210435

2. Sible IJ, Yew B, Dutt S, Bangen KJ, Li Y, Nation DA. Visit-to-visit blood pressure variability and regional cerebral perfusion decline in older adults. Neurobiol Aging. Sep 2021;105:57–63. doi:10.1016/j.neurobiolaging.2021.04.009

3. Sible IJ, Yew B, Dutt S, et al. Selective vulnerability of medial temporal regions to short-term blood pressure variability and cerebral hypoperfusion in older adults. Neuroimage Rep. Mar 2022;2(1)doi:10.1016/j.ynirp.2022.100080

4. Sible IJ, Nation DA. Blood Pressure Variability and Cerebral Perfusion Decline: A Post Hoc Analysis of the SPRINT MIND Trial. Journal of the American Heart Association. 2023/06/20 2023;12(12):e029797. doi:10.1161/JAHA.123.029797

5. Sible IJ, Jang JY, Dutt S, et al. Older Adults With Higher Blood Pressure Variability Exhibit Cerebrovascular Reactivity Deficits. American Journal of Hypertension. 2023;36(1):63–68. doi:10.1093/ajh/hpac108

6. Lohman T, Sible I, Kapoor A, et al. Blood Pressure Variability, Central Autonomic Network Dysfunction, and Cerebral Small-Vessel Disease in APOE4 Carriers. Journal of the American Heart Association. 2024/05/07 2024;13(9):e034116. doi:10.1161/JAHA.123.034116

7. Tully PJ, Yano Y, Launer LJ, et al. Association Between Blood Pressure Variability and Cerebral Small-Vessel Disease: A Systematic Review and Meta-Analysis. Journal of the American Heart Association. 2020/01/07 2020;9(1):e013841. doi:10.1161/JAHA.119.013841

8. Sible IJ, Nation DA, Weiner M, et al. Visit-to-Visit Blood Pressure Variability and Longitudinal Tau Accumulation in Older Adults. Hypertension. 2022;79(3):629–637. doi:10.1161/HYPERTENSIONAHA.121.18479

9. Sible IJ, Nation DA. Blood pressure variability and medial temporal atrophy in apolipoprotein L4 carriers. Brain Imaging Behav. Apr 2022;16(2):792–801. doi:10.1007/s11682-021-00553-1

10. Lohman T, Sible I, Engstrom AC, et al. Beat-to-beat blood pressure variability, hippocampal atrophy, and memory impairment in older adults. GeroScience. 2024/08/05 2024; doi:10.1007/s11357-024-01303-z

11. Asmuje NF, Mat S, Goh CH, Myint PK, Tan MP. Increased Beat-to-Beat Blood Pressure Variability Is Associated With Impaired Cognitive Function. Am J Hypertens. Dec 8 2022;35(12):998–1005. doi:10.1093/ajh/hpac107

12. de Havenon A, Anadani M, Prabhakaran S, Wong KH, Yaghi S, Rost N. Increased Blood Pressure Variability and the Risk of Probable Dementia or Mild Cognitive Impairment: A Post Hoc Analysis of the SPRINT MIND Trial. Journal of the American Heart Association. 2021/09/21 2021;10(18):e022206. doi:10.1161/JAHA.121.022206

13. Ebinger J, Driver M, Huang T, et al. Blood pressure variability supersedes heart rate variability as a real-world measure of dementia risk. Scientific Reports. 01/22 2024;14doi:10.1038/s41598-024-52406-8

14. Yoo JE, Shin DW, Han K, et al. Blood Pressure Variability and the Risk of Dementia. Hypertension. 2020/04/01 2020;75(4):982–990. doi:10.1161/HYPERTENSIONAHA.119.14033

15. de Heus RAA, Olde Rikkert MGM, Tully PJ, Lawlor BA, Claassen J. Blood Pressure Variability and Progression of Clinical Alzheimer Disease. Hypertension. Nov 2019;74(5):1172–1180. doi:10.1161/hypertensionaha.119.13664

16. Rouch L, Cestac P, Sallerin B, et al. Visit-to-Visit Blood Pressure Variability Is Associated With Cognitive Decline and Incident Dementia: The S.AGES Cohort. Hypertension. Oct 2020;76(4):1280–1288. doi:10.1161/hypertensionaha.119.14553

17. de Heus RAA, Tzourio C, Lee EJL, et al. Association Between Blood Pressure Variability With Dementia and Cognitive Impairment: A Systematic Review and Meta-Analysis. Hypertension. 2021/11/01 2021;78(5):1478–1489. doi:10.1161/HYPERTENSIONAHA.121.17797

18. Guo H, Tan Y, Yao Z, Zhang Z, Yan J, Meng X. Effect of visit-to-visit blood pressure variability on mild cognitive impairment and probable dementia in hypertensive patients receiving standard and intensive blood pressure treatment. Front Cardiovasc Med. 2023;10:1166554. doi:10.3389/fcvm.2023.1166554

19. Jia P, Hao Z, Yiu K, Tsoi K. Causal Effects Between Blood Pressure Variability and Alzheimer’s Disease: A Two-Sample Mendelian Randomization Study. J Clin Hypertens (Greenwich). May 2025;27(5):e70066. doi:10.1111/jch.70066

20. Lohman T, Sible I, Ma Y, Pahlevan NM, Nation DA. Blood Pressure Variability and Alzheimer Disease Biomarkers. Hypertension. 0(0)doi:10.1161/HYPERTENSIONAHA.126.26768

21. Sible IJ, Nation DA. Long-Term Blood Pressure Variability Across the Clinical and Biomarker Spectrum of Alzheimer’s Disease. J Alzheimers Dis. 2020;77(4):1655–1669. doi:10.3233/jad-200221

22. Sible IJ, Nation DA. Blood Pressure Variability and Plasma Alzheimer’s Disease Biomarkers in the SPRINT Trial. J Alzheimers Dis. 2024;97(4):1851–1860. doi:10.3233/jad-230930

23. Lattanzi S, Brigo F, Vernieri F, Silvestrini M. Visit-to-visit variability in blood pressure and Alzheimer’s disease. J Clin Hypertens (Greenwich). May 2018;20(5):918–924. doi:10.1111/jch.13290

24. Mancia G. Visit-to-Visit Blood Pressure Variability. Hypertension. 2016/07/01 2016;68(1):32–33. doi:10.1161/HYPERTENSIONAHA.116.07139

25. Gratz I, Deal E, Spitz F, et al. Continuous Non-invasive finger cuff CareTaker® comparable to invasive intra-arterial pressure in patients undergoing major intra-abdominal surgery. BMC Anesthesiol. Mar 21 2017;17(1):48. doi:10.1186/s12871-017-0337-z

26. Kwon Y, Stafford PL, Baruch MC, et al. Tracking of the beat-to-beat blood pressure changes by the Caretaker physiological monitor against invasive central aortic measurement. Blood Press Monit. Feb 1 2022;27(1):70–76. doi:10.1097/mbp.0000000000000568

27. Ooi JH, Lim R, Seng H, et al. Non-invasive parameters of autonomic function using beat-to-beat cardiovascular variations and arterial stiffness in hypertensive individuals: a systematic review. Biomed Eng Online. Feb 20 2024;23(1):23. doi:10.1186/s12938-024-01202-6

28. Lohman T, Sible IJ, Shenasa F, et al. Reliability of beat-to-beat blood pressure variability in older adults. Scientific Reports. 2024/08/30 2024;14(1):20197. doi:10.1038/s41598-024-71183-y

29. Sible IJ, Yew B, Jang JY, et al. Blood pressure variability and plasma Alzheimer’s disease biomarkers in older adults. Sci Rep. Oct 13 2022;12(1):17197. doi:10.1038/s41598-022-20627-4

30. Ashton NJ, Brum WS, Di Molfetta G, et al. Diagnostic Accuracy of a Plasma Phosphorylated Tau 217 Immunoassay for Alzheimer Disease Pathology. JAMA Neurology. 2024;81(3):255–263. doi:10.1001/jamaneurol.2023.5319

31. Jack CR, Jr., Andrews JS, Beach TG, et al. Revised criteria for diagnosis and staging of Alzheimer’s disease: Alzheimer’s Association Workgroup. Alzheimers Dement. Aug 2024;20(8):5143–5169. doi:10.1002/alz.13859

32. Khalafi M, Dartora WJ, McIntire LBJ, et al. Diagnostic accuracy of phosphorylated tau217 in detecting Alzheimer’s disease pathology among cognitively impaired and unimpaired: A systematic review and meta-analysis. Alzheimers Dement. Feb 2025;21(2):e14458. doi:10.1002/alz.14458

33. Ally M, Sugarman MA, Zetterberg H, et al. Cross-sectional and longitudinal evaluation of plasma glial fibrillary acidic protein to detect and predict clinical syndromes of Alzheimer’s disease. Alzheimer’s & Dementia: Diagnosis, Assessment & Disease Monitoring. 2023/10/01 2023;15(4):e12492. doi:10.1002/dad2.12492

34. Baiardi S, Quadalti C, Mammana A, et al. Diagnostic value of plasma p-tau181, NfL, and GFAP in a clinical setting cohort of prevalent neurodegenerative dementias. Alzheimers Res Ther. Oct 12 2022;14(1):153. doi:10.1186/s13195-022-01093-6

35. Bellaver B, Povala G, Ferreira PCL, et al. Plasma GFAP for populational enrichment of clinical trials in preclinical Alzheimer’s disease. Alzheimers Dement. May 2025;21(5):e70209. doi:10.1002/alz.70209

36. Benedet AL, Milà-Alomà M, Vrillon A, et al. Differences Between Plasma and Cerebrospinal Fluid Glial Fibrillary Acidic Protein Levels Across the Alzheimer Disease Continuum. JAMA Neurology. 2021;78(12):1471–1483. doi:10.1001/jamaneurol.2021.3671

37. Zou Y, Wang Y, Ma X, et al. CSF and blood glial fibrillary acidic protein for the diagnosis of Alzheimer’s disease: A systematic review and meta-analysis. Ageing Research Reviews. 2024/11/01/2024;101:102485. doi:10.1016/j.arr.2024.102485

38. Varma VR, An Y, Kac PR, et al. Longitudinal progression of blood biomarkers reveals a key role of reactive astrocytosis in preclinical Alzheimer’s disease. Med. 2025/06/09/2025:100724. doi:10.1016/j.medj.2025.100724

39. Pereira JB, Janelidze S, Smith R, et al. Plasma GFAP is an early marker of amyloid-β but not tau pathology in Alzheimer’s disease. Brain. 2021;144(11):3505–3516. doi:10.1093/brain/awab223

40. Mena L, Pintos S, Queipo NV, Aizpúrua JA, Maestre G, Sulbarán T. A reliable index for the prognostic significance of blood pressure variability. J Hypertens. Mar 2005;23(3):505–11. doi:10.1097/01.hjh.0000160205.81652.5a

41. Del Giorno R, Balestra L, Heiniger PS, Gabutti L. Blood pressure variability with different measurement methods: Reliability and predictors. A proof of concept cross sectional study in elderly hypertensive hospitalized patients. Medicine (Baltimore). Jul 2019;98(28):e16347. doi:10.1097/md.0000000000016347

42. Hixson JE, Vernier DT. Restriction isotyping of human apolipoprotein E by gene amplification and cleavage with HhaI. J Lipid Res. Mar 1990;31(3):545–8.

43. Doecke JD, Pérez-Grijalba V, Fandos N, et al. Total Aβ(42)/Aβ(40) ratio in plasma predicts amyloid-PET status, independent of clinical AD diagnosis. Neurology. Apr 14 2020;94(15):e1580–e1591. doi:10.1212/wnl.0000000000009240

44. R Core Team. R: A language and environment for statistical computing.: Foundation for Statistical Computing, Vienna, Austria.; 2022.

45. Benjamini Y, Hochberg Y. Controlling the False Discovery Rate: A Practical and Powerful Approach to Multiple Testing. Journal of the Royal Statistical Society: Series B (Methodological). 1995/01/01 1995;57(1):289–300. doi:10.1111/j.2517-6161.1995.tb02031.x

46. Kwon Y, Stafford PL, Enfield K, Mazimba S, Baruch MC. Continuous Noninvasive Blood Pressure Monitoring of Beat-By-Beat Blood Pressure and Heart Rate Using Caretaker Compared With Invasive Arterial Catheter in the Intensive Care Unit. Journal of Cardiothoracic and Vascular Anesthesia. 2022/07/01/2022;36(7):2012–2021. doi:10.1053/j.jvca.2021.09.042

47. Nation DA, Edland SD, Bondi MW, et al. Pulse pressure is associated with Alzheimer biomarkers in cognitively normal older adults. Neurology. Dec 3 2013;81(23):2024–7. doi:10.1212/01.wnl.0000436935.47657.78

48. Nation DA, Edmonds EC, Bangen KJ, et al. Pulse Pressure in Relation to Tau-Mediated Neurodegeneration, Cerebral Amyloidosis, and Progression to Dementia in Very Old Adults. JAMA Neurology. 2015;72(5):546–553. doi:10.1001/jamaneurol.2014.4477

49. Cooper LL, O’Donnell A, Beiser AS, et al. Association of Aortic Stiffness and Pressure Pulsatility With Global Amyloid-β and Regional Tau Burden Among Framingham Heart Study Participants Without Dementia. JAMA Neurology. 2022;79(7):710–719. doi:10.1001/jamaneurol.2022.1261

50. Bos I, Vos SJB, Schindler SE, et al. Vascular risk factors are associated with longitudinal changes in cerebrospinal fluid tau markers and cognition in preclinical Alzheimer’s disease. Alzheimers Dement. Sep 2019;15(9):1149–1159. doi:10.1016/j.jalz.2019.04.015

51. Laosiripisan J, Haley AP, Tanaka H. Steady State vs. Pulsatile Blood Pressure Component and Regional Cerebral Perfusion. Am J Hypertens. Nov 1 2017;30(11):1100–1105. doi:10.1093/ajh/hpx108

52. Marina N, Christie IN, Korsak A, et al. Astrocytes monitor cerebral perfusion and control systemic circulation to maintain brain blood flow. Nat Commun. Jan 9 2020;11(1):131. doi:10.1038/s41467-019-13956-y

53. Haidey JN, Peringod G, Institoris A, et al. Astrocytes regulate ultra-slow arteriole oscillations via stretch-mediated TRPV4-COX-1 feedback. Cell Rep. Aug 3 2021;36(5):109405. doi:10.1016/j.celrep.2021.109405

54. Belko Parkel K, Kuhanec D, Gržina Ž T, Haque Chowdhury H, Zorec R, Kreft M. Brief mechanical pulses induce sustained intracellular L-lactate production in astrocytes. Front Cell Neurosci. 2026;20:1830831. doi:10.3389/fncel.2026.1830831

55. Strat AN, Ahmed A, Viapiano M, Ganapathy PS. Astrocyte Mechanobiology: Linking Biomechanical Forces to Biochemical Signaling in the Central Nervous System. Glia. Jul 2026;74(7):e70167. doi:10.1002/glia.70167

56. Diaz JR, Kim KJ, Brands MW, Filosa JA. Augmented astrocyte microdomain Ca(2+) dynamics and parenchymal arteriole tone in angiotensin II-infused hypertensive mice. Glia. Mar 2019;67(3):551–565. doi:10.1002/glia.23564

57. Filosa JA, Iddings JA. Astrocyte regulation of cerebral vascular tone. Am J Physiol Heart Circ Physiol. Sep 1 2013;305(5):H609–19. doi:10.1152/ajpheart.00359.2013

58. Filosa JA, Morrison HW, Iddings JA, D. W, Kim KJ. Beyond neurovascular coupling, role of astrocytes in the regulation of vascular tone. Neuroscience. May 26 2016;323:96–109. doi:10.1016/j.neuroscience.2015.03.064

59. Kim KJ, Iddings JA, Stern JE, et al. Astrocyte Contributions to Flow/Pressure-Evoked Parenchymal Arteriole Vasoconstriction. The Journal of Neuroscience. 2015;35(21):8245. doi:10.1523/JNEUROSCI.4486-14.2015

60. Price BR, Johnson LA, Norris CM. Reactive astrocytes: The nexus of pathological and clinical hallmarks of Alzheimer’s disease. Ageing Research Reviews. 2021/07/01/2021;68:101335. doi:10.1016/j.arr.2021.101335

61. Bellaver B, Povala G, Ferreira PCL, et al. Astrocyte reactivity influences amyloid-β effects on tau pathology in preclinical Alzheimer’s disease. Nature Medicine. 2023/07/01 2023;29(7):1775–1781. doi:10.1038/s41591-023-02380-x

62. Kim JS, Yi D, Byun MS, et al. Plasma GFAP as a mediator and moderator of Aβ, tau, and neurodegeneration in Alzheimer’s disease. npj Dementia. 2026/01/09 2026;2(1):3. doi:10.1038/s44400-025-00051-x

63. Allwright M, Mundell HD, McCorkindale AN, et al. Ranking the risk factors for Alzheimer’s disease; findings from the UK Biobank study. Aging Brain. 2023/01/01/2023;3:100081. doi:10.1016/j.nbas.2023.100081

64. Doecke JD, Chenna A, Lo M, et al. Comparing plasma Aβ42/40 and pTau217 to predict Amyloid-β status across the Alzheimer’s disease continuum. Alzheimers Dement. © 2024 The Alzheimer’s Association. Alzheimer’s & Dementia published by Wiley Periodicals LLC on behalf of Alzheimer’s Association.; 2024. vol. Suppl 2.

65. Wennström M, Janelidze S, Nilsson KPR, et al. Cellular localization of p-tau217 in brain and its association with p-tau217 plasma levels. Acta Neuropathol Commun. Jan 6 2022;10(1):3. doi:10.1186/s40478-021-01307-2

66. Kang H, Yoo H, Lee J, et al. Plasma phosphorylated tau 217 and amyloidl1Jβ 42/40 for amyloid risk in subgroups. Alzheimers Res Ther. Aug 7 2025;17(1):184. doi:10.1186/s13195-025-01826-3

67. Cánovas R, Cox T, Doré V, et al. When Does Alzheimer’s Disease Start? Plasma Aβ42/40 Assays Show Steep Changes at Aβ-PET Centiloid 15, Mean Age of 66LYears. Ann Neurol. May 2026;99(5):1327–1342. doi:10.1002/ana.78163

68. Ashton NJ, Janelidze S, Mattsson-Carlgren N, et al. Differential roles of Aβ42/40, p-tau231 and p-tau217 for Alzheimer’s trial selection and disease monitoring. Nat Med. Dec 2022;28(12):2555–2562. doi:10.1038/s41591-022-02074-w

69. Yu L, Boyle PA, Janelidze S, et al. Plasma p-tau181 and p-tau217 in discriminating PART, AD and other key neuropathologies in older adults. Acta Neuropathol. Jul 2023;146(1):1–11. doi:10.1007/s00401-023-02570-4

